# Feature harmonization reveals that apparent cross-cohort differences in tau-positivity prediction reflect variable availability and cohort stage structure

**DOI:** 10.64898/2026.10.16.26365770

**Authors:** Avinay Bhat, Mattia Cestari, Chin-Tu Chen, Bonnie T. Fleming, George T. Fleming, the Alzheimer’s Disease Neuroimaging Initiative

## Abstract

**Background:** Cross-cohort machine-learning studies of tau positivity are increasingly used to assess generalizability of Alzheimer’s disease biomarker models, but apparent differences between cohorts may reflect clinical severity structure and cohort stage composition rather than genuine biological divergence.

**Methods:** We evaluated this problem across subject-level cohorts from the Alzheimer’s Disease Neuroimaging Initiative (ADNI), the Open Access Series of Imaging Studies (OASIS3), and the National Alzheimer’s Coordinating Center (NACC) using harmonized tabular predictors and a feature-block ablation framework. For ADNI and OASIS3, we performed matched-feature analyses restricted to shared harmonized variables (age, APOE ***ε***4, education, sex, the CDR^®^ Dementia Staging Instrument sum-of-boxes score (CDR-SB), and the Functional Activities Questionnaire (FAQ)). Models were trained with five-fold stratified cross-validation using balanced logistic regression as the primary classifier. Random forests and extra-trees classifiers were evaluated as robustness checks and are reported in supplementary analyses.

**Results:** In the primary matched-feature comparison, reference areas under the receiver operating characteristic curve (AUCs) were 0.851 for ADNI tau90, 0.860 for ADNI tau180, and 0.933 for OASIS3 tau180. Severity-only models retained 86–99% of reference performance, whereas demographic/genetic-only models retained 85–87%; bootstrap confidence intervals showed that the reference-minus-demo contrast remained positive across all primary cohorts. Before feature matching, OASIS3 showed substantially weaker demographic-only prediction (AUC ***≈***0.54), attributable to missing APOE and age variables. After harmonization with a matched feature set including age, the pre-harmonization demographic gap was no longer attributable to feature availability; the residual difference was most consistent with cohort disease-stage composition asymmetry, with CDR-SB **=** 0 prevalence of 84.5–88.7% in OASIS3 versus 49.3–51.7% in ADNI.

**Conclusions:** The original large ADNI–OASIS3 performance gap was largely a feature-availability artifact; after strict harmonization including age-field repair, the absolute gap narrowed and the signed ADNI-minus-OASIS3 difference reversed, with the residual difference most consistent with cohort disease-stage composition asymmetry. Clinical severity remains an important predictor across all cohorts, but the unique contributions of severity and demographic/genetic information depend strongly on stage composition. Cross-cohort biomarker modeling should account for both feature-set completeness and disease-stage composition heterogeneity, not only headline performance metrics.

## 1 Introduction

Tau positron emission tomography (PET) has become central to Alzheimer’s disease (AD) research, providing in vivo quantification of neurofibrillary tangle burden that correlates with disease stage and clinical outcomes [1, 2]. Machine-learning models that predict tau-PET positivity from more accessible clinical and genetic variables are attracting growing interest, both as potential screening tools and as a means of characterizing the information content of different predictor domains [3–5]. Recent work has demonstrated that plasma biomarkers, particularly phosphorylated tau-217, carry substantial predictive signal for tau-PET burden [3, 6], while clinical and demographic variables have been used to build risk scores in single-cohort settings [5].

A key challenge for these models is cross-cohort generalizability. Major AD research cohorts—including the Alzheimer’s Disease Neuroimaging Initiative (ADNI), the Open Access Series of Imaging Studies (OASIS), and the National Alzheimer’s Coordinating Center (NACC)—differ in enrollment criteria, clinical populations, available variables, and endpoint definitions. Prior work has addressed harmonization of PET imaging data across tracers and scanners [7, 8] and has developed ontology-based approaches for aligning clinical data elements across cohorts [9]. Cross-cohort evaluation studies have also demonstrated that cohort differences can materially affect model performance in dementia prediction [10]. Most recently, Lin et al. classified tau status in amyloid-positive individuals from structural MRI, amyloid-PET and demographic features, training in ADNI and validating externally in OASIS-3 and SCAN [11], and multi-cohort models built on clinically accessible features have been validated in NACC [12]. Neither line of work, however, decomposes how clinical severity and demographic features contribute to cross-cohort performance differences. More broadly, most cross-cohort comparisons report headline performance metrics without systematically decomposing *what* the models are learning.

This decomposition matters because tau positivity is not biologically independent of clinical severity. Clinical dementia rating scales, functional assessments, and cognitive screening scores are themselves downstream consequences of the neurodegenerative process that drives tau accumulation. Models that include these severity measures may therefore be predicting a correlate of disease stage rather than identifying upstream biological risk. Recent work on tau–clinical mismatch has shown that roughly half of amyloid-positive individuals display “canonical” concordance between tau burden and clinical severity, while the remainder are classified as either resilient or vulnerable [13]. This suggests that the tau–severity relationship, while strong on average, is not deterministic, and that disentangling severity-driven from biology-driven signal is scientifically meaningful.

The present study addresses this gap using a feature-block ablation framework applied identically across ADNI, OASIS3, and NACC. We first quantified how much predictive performance could be attributed to severity versus demographic/genetic feature blocks within each cohort. We then performed a matched-feature rerun restricted to variables genuinely shared across cohorts after harmonization, feature auditing, and age-field repair. Our goal was not to build an optimal tau classifier, but to answer a methodological question: when cohorts appear to differ in tau-positivity prediction, how much of that difference survives after the feature space is matched and severity signal is explicitly decomposed?

## 2 Methods

### 2.1 Study design

Data used in the preparation of this article were obtained from the Alzheimer’s Disease Neuroimaging Initiative (ADNI) database (https://adni.loni.usc.edu/). ADNI was launched in 2003 as a public-private partnership, led by Principal Investigator Michael W. Weiner, MD. The original goal of ADNI was to test whether serial magnetic resonance imaging (MRI), positron emission tomography (PET), other biological markers, and clinical and neuropsychological assessment could be combined to measure the progression of mild cognitive impairment (MCI) and early Alzheimer’s disease (AD). Current goals include validating biomarkers for clinical trials, improving the generalizability of ADNI data by increasing diversity in the participant cohort, and providing data concerning the diagnosis and progression of Alzheimer’s disease to the scientific community. For up-to-date information, see https://adni.loni.usc.edu/. ADNI data used in this study were downloaded from the ADNI repository on March 6th 2026, and this download date is reported here in accordance with the ADNI Data Use Agreement.

We conducted a retrospective cross-cohort analysis using subject-level data from ADNI, OASIS3 [14], and NACC. The primary endpoint family was binary tau positivity in ADNI and OASIS3. NACC was included as a supplementary endpoint-family anchor using a strict A+/T+ versus A*−*/T*−* definition. The study comprised three sequential analyses: (i) within-cohort feature-block ablation with all available harmonized features, (ii) matched-feature rerun across ADNI and OASIS3 restricted to shared variables including restored age, and (iii) supplementary anchoring against NACC. An overview of the full workflow is shown in Fig. 1.

**Fig. 1.**
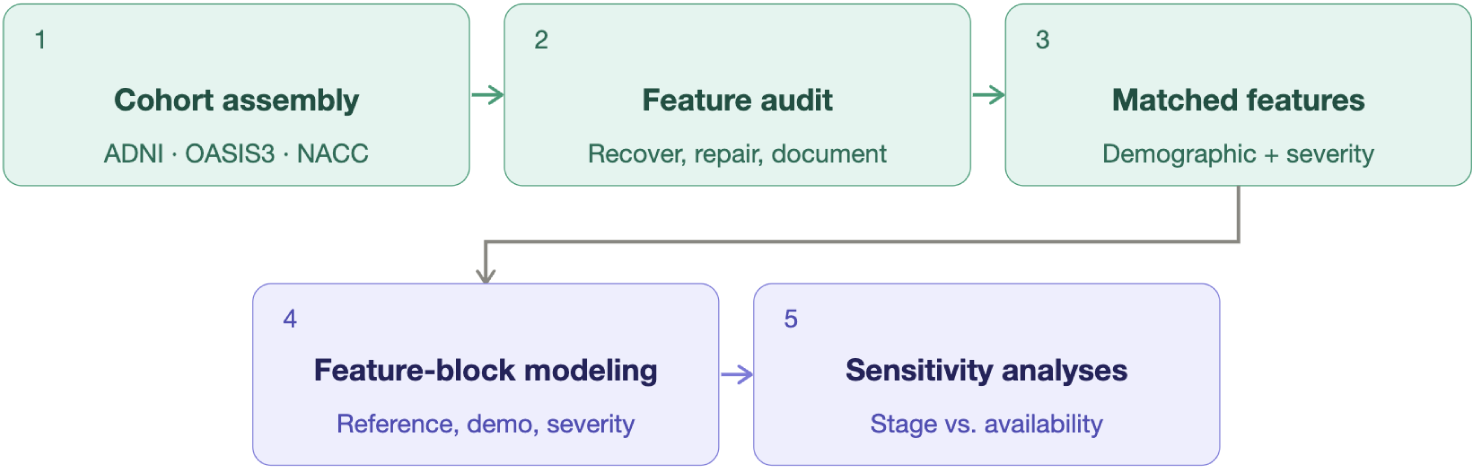
Study overview and harmonization workflow. Subject-level cohorts were assembled from ADNI, OASIS3, and NACC, followed by explicit feature auditing and harmonization. The audit recovered age and APOE variables in OASIS3, documented unavailable variables, and repaired the ADNI age field using PTDEMOG-derived age and elapsed time. The primary ADNI–OASIS3 analysis used a matched feature set spanning demographic/genetic predictors and clinical-severity predictors. Feature-block ablation was evaluated using balanced logistic regression as the primary classifier, with random forest and extra-trees classifiers used as robustness checks. Sensitivity analyses assessed CDR-SB= 0 subgroup structure, OASIS3 tau-threshold robustness, within-OASIS3 continuous tauopathy SUVR, and diagnosis-stratified feature-block performance to distinguish feature-availability artifacts from residual cohort stage-composition differences.

### 2.2 Cohorts and endpoints

#### ADNI tau cohorts

Two ADNI subject-level tau-positivity cohorts were derived from previously constructed modality-matched tables with 90-day and 180-day matching windows. The ADNI tau90 cohort included 438 subjects (78 tau positive; 17.8%), and the ADNI tau180 cohort included 547 subjects (84 tau positive; 15.4%). Tau positivity labels were taken from finalized cohort tables in which positivity had been determined using established flortaucipir meta-temporal region-of-interest thresholds applied during cohort construction.

#### OASIS3 tau cohorts

OASIS3 90-day and 180-day subject-level cohorts were constructed using the same modality-matching logic. After one-row-per-subject selection favoring visits with the closest tau–visit temporal match and lowest feature missingness, the OASIS3 tau90 cohort included 142 subjects (12 tau positive; 8.5%) and the OASIS3 tau180 cohort included 368 subjects (45 tau positive; 12.2%).

Because OASIS3 does not distribute a binary tau-positivity label, tau positivity was derived by fitting a two-component Gaussian mixture model (GMM) to the tauopathy standardized uptake value ratio (SUVR) summary variable distributed with OASIS-3 (Tauopathy: the mean partial-volume-corrected SUVR of the amygdala, entorhinal cortex, inferior temporal cortex and lateral occipital cortex, with a cerebellar cortex reference region [15, 16]), with 10 random initializations and a fixed random seed. The positivity threshold was set at the midpoint of the two fitted component means. The OASIS-3 imaging data dictionary also specifies a tau-positivity cutoff of 1.22 for this measure [16]. In our cohorts, however, that cutoff classified 46.5% of OASIS3 tau180 participants as tau-positive (171/368; 45.1%, 64/142, in tau90), whereas only 29.8% of OASIS3 tau180 participants with amyloid PET (93/312) were amyloid-positive; among tau-positive participants with amyloid PET, 51.1% (72/141) were amyloid-negative at the published cutoff, compared with 5.6% (2/36) at the GMM midpoint. Because the ADNI endpoint targets AD-type (meta-temporal) tau positivity, we retained the GMM midpoint as the primary OASIS3 definition and report results at the published cutoff in Supplementary Table S5. Amyloid status for this comparison was taken from the matched amyloid PET scan using the OASIS-3 cutoffs for partial-volume-corrected mean cortical SUVR (Pittsburgh compound B 1.42; florbetapir 1.19) [16]. The GMM midpoint nonetheless introduces a methodological asymmetry relative to the ADNI threshold, which we address in the limitations. Because the OASIS3 tau90 cohort contained only 12 positive cases (approximately 2–3 per cross-validation fold), it was treated as supplementary throughout.

#### NACC strict A/T cohort

As a supplementary sensitivity anchor, we used a strict binary NACC endpoint contrasting A+/T+ against A*−*/T*−*, excluding intermediate cases. The main strict cohort included 413 subjects (109 A+/T+, 304 A*−*/T*−*). Amyloid positivity was defined using cohort-provided PET-based amyloid status, and tau positivity was defined using cohort-provided tau-PET status. Two tau tracers were represented in this cohort (flortaucipir, *n* = 296; MK-6240, *n* = 117); accordingly, tau tracer identity was included as a feature in the reference and demographic-only NACC models. In addition to the harmonized variables listed in Section 2.3, the NACC demographic block also included race, Hispanic ethnicity and handedness, and the NACC severity block also included animal and vegetable category fluency. Because this endpoint family differs from ADNI/OASIS tau positivity, NACC results were treated as confirmatory and were not directly pooled with the primary analysis. We note that some ADNI sites contribute data to NACC; however, because NACC was used as a different-endpoint sensitivity anchor rather than a direct replication target, and because the feature sets and endpoint definitions differ, potential subject overlap does not invalidate the comparison.

### 2.3 Feature harmonization

Feature harmonization targeted demographic/genetic and clinical-severity domains. Across all cohorts, the following harmonized variables were considered when available: age, sex, education (years), APOE *ε*4 allele count, APOE *ε*4 carrier status (binary), CDR^®^ Dementia Staging Instrument global score and sum of boxes (CDR-SB), Functional Activities Questionnaire (FAQ) total score, and Montreal Cognitive Assessment (MoCA) total score.

Variable recovery required cohort-specific column identification. In ADNI, demographic variables were drawn from a master visits table and merged to subject-level cohort tables by subject identifier and visit date. APOE genotype was parsed from a two-allele string (e.g., “3/4”) to an *ε*4 count. Sex was harmonized from numeric coding (1/2) to categorical (M/F). Age was derived from PTDEMOG-based age at baseline plus elapsed days from baseline to tau-PET visit, and validated to lie within the plausible range 40–100 years; any field containing epoch-format timestamps rather than chronological years was flagged and repaired in the upstream pipeline. In OASIS3, an analogous column-matching procedure was applied, with APOE parsed from a genotype string column and CDR-SB mapped from a CDRSUM column.

#### Feature audit

A dedicated feature audit was performed on the OASIS3 raw and subject-level files to determine whether apparently missing variables (age, APOE, CDR global, MoCA) were genuinely absent or merely unrecovered due to naming mismatches. The audit showed that age and APOE were recoverable, whereas MoCA and CDR global were absent from the current OASIS3 clinical master extract. After recovery, OASIS3 subject-level tables contained age (368/368 non-missing in tau180), APOE *ε*4 count (367/368), education (368/368), sex (368/368), CDR-SB (368/368), and FAQ (367/368).

#### Matched-feature restriction

The final matched-feature primary analysis used variables consistently present across all primary cohorts: age, APOE *ε*4 count, APOE *ε*4 carrier status, education, and sex (demographic/genetic block); CDR-SB and FAQ total (severity block). Age was successfully recovered in both ADNI cohorts following the PTDEMOG-based repair described above (tau90: 72.90*±*7.68 years; tau180: 72.78*±*7.87 years), and was present in both OASIS3 cohorts from the feature audit. The inclusion of age in both cohort families enabled a more complete matched-feature comparison than would otherwise be possible.

### 2.4 Subject-level table construction

All analyses were performed at the subject level using one row per subject. Where multiple source rows existed for a subject, the pipeline selected the best candidate by minimizing temporal distance to the tau-PET scan and maximizing feature completeness. For ADNI, this included merging the final tau cohort table with the ADNI master visits table and retaining the best available harmonized feature record per subject. For OASIS3, subject-level rows were selected using tau-window matching logic that prioritized temporal proximity and low feature missingness.

### 2.5 Model families and evaluation

Balanced logistic regression was used as the primary classifier across all main analyses (liblinear solver, L2 regularization, class-balanced weights, maximum 5000 iterations). This choice was prespecified to prioritize interpretability, stability in modest sample sizes, and clinical auditability of demographic and severity-domain effects. Random forest and extra-trees classifiers were evaluated as robustness checks using the same cross-validation splits and preprocessing pipeline; these results are reported in the supplementary materials. All tables and main-text figures, including the NACC results in Table 4, report the balanced-logistic-regression results only (random-forest and extra-trees results appear only in Additional file 1); best-performing model selection across classifier families was not used.

Evaluation used five-fold stratified cross-validation with a single fixed split (random seed 42). Preprocessing was performed inside each fold via a scikit-learn pipeline: numeric features received median imputation followed by standard scaling; categorical features received most-frequent imputation followed by one-hot encoding. For the initial all-available within-cohort feature-block analysis, features with *>*35% missingness were excluded prior to model fitting. In contrast, the matched-feature primary rerun used a prespecified shared feature set, including FAQ as a shared severity variable, with missing values handled by foldwise imputation inside the cross-validation pipeline.

Performance was summarized using mean cross-validated area under the receiver operating characteristic curve (AUC), average precision (AP), balanced accuracy, and F1 score. For the three primary matched-feature cohorts (ADNI tau90, ADNI tau180, and OASIS3 tau180), we estimated 95% bootstrap confidence intervals for AUCs and for the pairwise contrasts reference minus severity-only, reference minus demo-only, and reference minus minus-all-CDR using 1000 subject-level bootstrap resamples.

### 2.6 Feature-block ablation design

The feature-block ablation framework decomposed prediction into clinically interpretable blocks. For each cohort, we evaluated:

- **reference-combined:** all shared demographic/genetic plus severity features;
- **demo-only:** demographic/genetic features only (age, APOE, education, sex);
- **severity-only:** severity features only (CDR-SB, FAQ);
- **minus-all-CDR:** reference model after removing all CDR variables;
- **minus-FAQ:** reference model after removing FAQ.

The primary derived metrics were:

- severity retention fraction = AUC_severity_*/*AUC_reference_,
- demo retention fraction = AUC_demo_*/*AUC_reference_,
- ^•^ CDR block drop = AUC_reference_ *−* AUC_minus-all-CDR_,
- full-strip drop = AUC_reference_ *−* AUC_demo_.

### 2.7 Primary versus supplementary analyses

The primary table was restricted to ADNI tau90, ADNI tau180, and OASIS3 tau180—the cohorts forming the most interpretable matched-feature comparison. The supplementary table included OASIS3 tau90 (small positive count) and NACC strict A/T (different endpoint family).

### 2.8 Sensitivity analyses

Four additional sensitivity analyses were performed to evaluate the robustness of the stage-composition and OASIS3 threshold interpretations.

First, we repeated cohort summaries within the CDR-SB = 0 subgroup to assess whether the apparent cross-cohort stage asymmetry persisted among subjects without detectable functional impairment. Because the resulting tau-positive counts were very small in several cohorts, within-stage AUCs were interpreted cautiously and were not treated as primary performance estimates.

Second, because OASIS3 tau positivity was derived from a data-driven GMM threshold rather than an externally fixed flortaucipir cutoff, we performed a threshold-sensitivity analysis around the OASIS3 GMM midpoint. This analysis used demographic/genetic predictors only (age, APOE *ε*4 count, APOE *ε*4 carrier status, education, and sex) so that changes in demographic discriminability could be attributed to the tau-positivity threshold rather than to inclusion of clinical severity variables. Thresholds yielding fewer than 10 tau-positive subjects were flagged as unreliable. Results at the published OASIS-3 tauopathy cutoff (1.22) [16] are reported alongside the sweep.

Third, as an additional within-OASIS3 sensitivity analysis, we evaluated whether the feature-block pattern persisted when the harmonized tauopathy SUVR variable was modeled continuously rather than binarized at the GMM midpoint. Within OASIS3 tau90 and tau180, we fit ridge-regression models using the same matched feature blocks as in the binary analysis (reference-combined, demo-only, severity-only, minus-all-CDR, and minus-FAQ; in the matched OASIS3 feature set, minus-all-CDR is operationalized as removing CDR-SB). Model performance was assessed using target-stratified five-fold cross-validation, with continuous Tauopathy SUVR as the outcome and cross-validated *R*^2^ as the primary metric. This analysis was not used for direct ADNI–OASIS3 comparison because continuous SUVR values are not on a common scale across cohorts, tracers, preprocessing pipelines, and reference regions; it was used only to test whether the OASIS3 conclusions were dependent on the binary GMM threshold.

Fourth, to directly test the severity-entanglement hypothesis, we repeated the reference-combined and demo-only experiments within clinical diagnosis strata (CN, MCI, Dementia) defined by baseline diagnosis in ADNI (DX DIAGNOSIS: 1 = CN, 2 = MCI, 3 = Dementia) and OASIS3 (dx harmonized). Strata with fewer than 10 tau-positive or 10 tau-negative subjects were not estimated. The primary hypothesis was that severity features would be less informative—and potentially detrimental—in functionally intact (CN) subjects where CDR-SB is near-uniformly zero, and more informative in Dementia subjects where functional impairment varies substantially. Standardized logistic regression coefficients were extracted from the primary matched-feature reference-combined model to quantify the direction and relative magnitude of each feature’s contribution (Supplementary Table S6).

## 3 Results

### 3.1 Cohort characteristics

Table 1 summarizes the demographic and clinical characteristics of the analytic samples. CDR-SB = 0 prevalence is reported per cohort as a proxy for the fraction of cognitively unimpaired subjects, which is relevant for interpreting severity-driven prediction.

**Table 1.** Demographic and clinical characteristics of the ADNI and OASIS3 analytic samples. Values are mean *±* SD or *n* (%). Female percentages are calculated using the full cohort *N*. Percentages for APOE *ε*4 carrier status and CDR-SB = 0 are calculated among non-missing values for that variable. Age in ADNI cohorts was recovered via PTDEMOG-based derivation following repair of epoch-timestamp contamination in the upstream harmonized field. The two ADNI cohorts share substantial participant overlap (tau180 uses a wider matching window that subsumes most tau90 subjects), which explains their similar demographic distributions.

|  | ADNI tau90<br>(primary) | ADNI tau180<br>(primary) | OASIS3 tau180<br>(primary) | OASIS3 tau90<br>(supp.) |
| --- | --- | --- | --- | --- |
| $N$ | 438 | 547 | 368 | 142 |
| Tau pos., $n$ (%) | 78 (17.8) | 84 (15.4) | 45 (12.2) | 12 (8.5) |
| Female, $n$ (%) | 245 (55.9) | 319 (58.3) | 207 (56.2) | 82 (57.7) |
| Age, mean ± SD | 72.90 ± 7.68 | 72.78 ± 7.87 | 69.44 ± 8.41 | 68.91 ± 8.00 |
| Education (yrs), mean ± SD | 16.46 ± 2.38 | 16.41 ± 2.38 | 16.34 ± 2.40 | 16.33 ± 2.41 |
| APOE $\epsilon 4$+, $n$ (%) | 175 (40.8) | 209 (39.5) | 149 (40.6) | 57 (40.1) |
| CDR-SB, mean ± SD | 1.19 ± 1.93 | 1.10 ± 1.88 | 0.37 ± 1.11 | 0.29 ± 0.95 |
| CDR-SB = 0, $n$ (%) | 215 (49.3) | 282 (51.7) | 311 (84.5) | 126 (88.7) |
| FAQ total, mean ± SD | 2.69 ± 5.65 | 2.42 ± 5.45 | 3.00 ± 5.94 | 3.22 ± 6.34 |

**Table 2.** Effect of feature harmonization on the cross-cohort demo-only comparison (ADNI tau180 vs. OASIS3 tau180). The pre-harmonization comparison used all features available in each cohort separately; the post-harmonization comparison used only shared features including recovered age.

| Setting | ADNI tau180 | OASIS3 tau180 | Absolute gap |
| --- | --- | --- | --- |
| Demo-only AUC (pre-harmonization) | 0.742 | 0.538 | 0.204 |
| Demo features used (pre) | 4 variables | 2 variables | — |
| Demo-only AUC (post-harmonization) | 0.741 | 0.793 | 0.052* |
| Demo features used (post) | 5 variables | 5 variables | — |

Pre-harmonization ADNI demo features: age, APOE $\epsilon$ 4 count, education, sex.
Pre-harmonization OASIS3 demo features: education, sex only.
Pre-harmonization values were recomputed with the pre-audit feature sets on the final, age-repaired subject tables.
Post-harmonization demo features in both cohorts: age, APOE $\epsilon$ 4 count, APOE $\epsilon$ 4 carrier, education, sex.
\*The signed ADNI-minus-OASIS3 difference reverses after harmonization ( $0.741 - 0.793 = -0.052$ ): OASIS3 demo-only AUC exceeds ADNI after feature matching, reflecting OASIS3’s earlier disease-stage composition (Section 3.3).

### 3.2 Feature availability before and after audit

As summarized in Fig. 1, the cross-cohort pipeline included an explicit OASIS3 feature-audit step before the matched-feature rerun. Prior to this audit and variable recovery, the OASIS3 subject-level tables lacked age, APOE *ε*4 count, APOE *ε*4 carrier status, CDR global, and MoCA. A pre-audit diagnostic feature-block ablation using all features available in each cohort showed OASIS3 tau180 demo-only AUC of 0.538 with only education and sex as predictors, compared to ADNI tau180 demo-only AUC of 0.742 with age, APOE, education, and sex.

Following the OASIS3 feature audit and ADNI age-field repair, both cohort families were compared using an identical shared demographic block comprising age, APOE *ε*4 count, APOE *ε*4 carrier status, years of education, and sex. Under this matched-feature setting, the demo-only AUC was 0.743 (95% CI 0.678–0.802) for ADNI tau90, 0.741 (95% CI 0.686–0.805) for ADNI tau180, and 0.793 (95% CI 0.729–0.874) for OASIS3 tau180. The residual cross-cohort difference was modest and persisted after harmonization, arguing against feature availability as the sole explanation and instead suggesting that cohort disease-stage composition contributes to the remaining difference, as detailed in Section 3.3.

### 3.3 Cohort composition and disease-stage structure

CDR-SB = 0 prevalence—the proportion of subjects with no detected functional impairment across any cognitive domain—differed markedly across cohorts (Fig. 3): 49.3% in ADNI tau90 (215/438), 51.7% in ADNI tau180 (282/547), 84.5% in OASIS3 tau180 (311/368), and 88.7% in OASIS3 tau90 (126/142). These differences indicate that the OASIS3 analytic samples were substantially earlier-stage and less severity-enriched than the corresponding ADNI cohorts. In such a setting, demographic and genetic predictors would be expected to retain more independent signal above severity, consistent with the observed pattern of severity and demo-only AUCs in OASIS3 tau180 (severity-only 0.799, 95% CI 0.727–0.885; demo-only 0.793, 95% CI 0.729–0.874).

### 3.4 CDR-SB=0 and OASIS3 sensitivity analyses

In a CDR-SB = 0 restricted sensitivity analysis, tau-positive prevalence fell sharply across all cohorts. Among subjects with CDR-SB = 0, tau-positive counts were 5/215 (2.3%) in ADNI tau90, 5/282 (1.8%) in ADNI tau180, 15/311 (4.8%) in OASIS3 tau180, and 4/126 (3.2%) in OASIS3 tau90. The very small number of tau-positive ADNI subjects within this stratum precluded reliable within-stage AUC estimation. Nevertheless, the base-rate collapse supports the interpretation that disease-stage composition strongly structures the apparent cross-cohort prediction problem.

OASIS3 threshold-sensitivity analysis further supported the treatment of OASIS3 tau180 as the primary OASIS3 cohort and OASIS3 tau90 as supplementary. At the OASIS3 tau180 GMM midpoint threshold based on the harmonized tauopathy variable (SUVR = 1.536), demographic/genetic-only balanced logistic regression yielded AUC = 0.793 across all 368 subjects with 45 tau-positive cases. Across reliable thresholds in the *±*30% sweep around the midpoint, tau180 AUCs ranged from 0.677 to 0.800, indicating that the demographic/genetic signal was not driven by a single brittle cut point. At the published OASIS-3 cutoff (1.22), demographic/genetic-only AUC was 0.683 in tau180 (171 tau-positive) and 0.665 in tau90 (64 tau-positive). Most participants classified as tau-positive at this cutoff had CDR-SB = 0 (74.3% in tau180, versus 33.3% at the GMM midpoint), and severity-only AUC fell to 0.577, consistent with this lower cutoff labelling many clinically unimpaired, amyloid-negative participants as tau-positive. In OASIS3 tau90, the GMM midpoint threshold (SUVR = 1.596) yielded 12 tau-positive subjects and AUC = 0.731; however, because the cohort remained small, with only approximately 2–3 positive cases per cross-validation fold, OASIS3 tau90 was retained as a supplementary analysis. Complete threshold-sweep results are provided in Supplementary Fig. S1 and Supplementary Table S5.

A within-OASIS3 continuous-SUVR sensitivity analysis supported the conclusion that the OASIS3 findings were not solely an artifact of the GMM binary threshold. Using continuous Tauopathy SUVR as the outcome, ridge-regression reference-combined models achieved cross-validated *R*^2^ = 0.481 in OASIS3 tau180 and *R*^2^ = 0.488 in OASIS3 tau90 (Supplementary Table S7; Supplementary Fig. S3). Severity-only models retained most of this continuous-burden signal (*R*^2^ = 0.424 in tau180 and 0.466 in tau90), whereas demo-only models explained substantially less variance (*R*^2^ = 0.132 in tau180 and 0.096 in tau90). Removing CDR-SB reduced performance sharply in both tau180 (*R*^2^ = 0.481 to 0.241) and tau90 (*R*^2^ = 0.488 to 0.155), whereas removing FAQ had little effect in either cohort (tau180: *R*^2^ = 0.481 to 0.486; tau90: *R*^2^ = 0.488 to 0.481). Thus, even without binary thresholding, OASIS3 tau burden was strongly linked to clinical-severity structure, particularly CDR-SB.

### 3.5 Matched-feature primary analysis

The central results of the matched-feature analysis are presented in Table 3 and Fig. 2. Reference-combined AUCs were 0.851 for ADNI tau90, 0.860 for ADNI tau180, and 0.933 for OASIS3 tau180. The notably higher OASIS3 tau180 reference AUC relative to both ADNI cohorts likely reflects the disease-stage composition asymmetry described in Section 3.3: in a predominantly clinically unimpaired sample, the combined feature set—particularly age and APOE—provides stronger discrimination than in a severity-enriched cohort where much of the predictive signal is already captured by clinical measures.

**Fig. 2.**
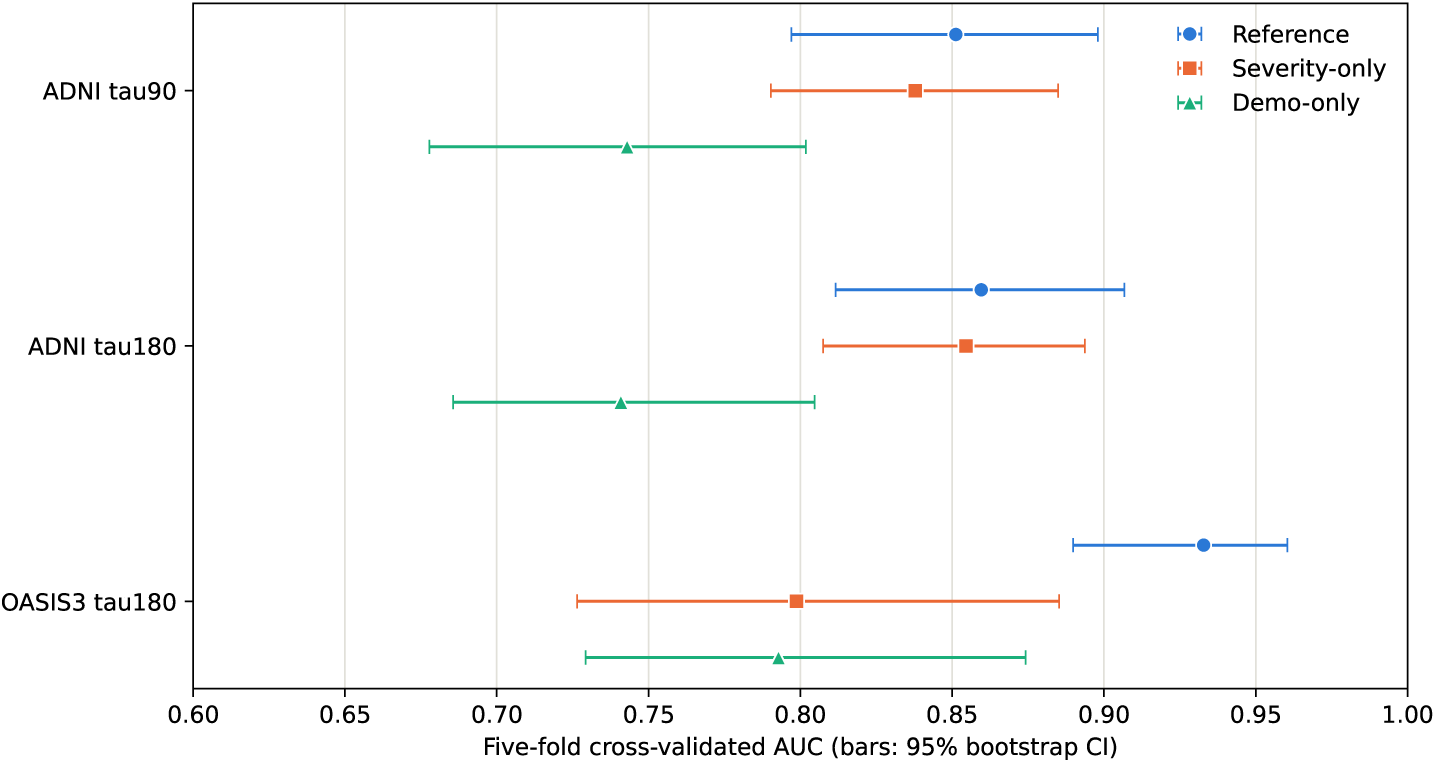
Cross-cohort decomposition of tau-positivity prediction by feature domain. Primary matched-feature analysis. Points show five-fold cross-validated AUC (Table 3) and horizontal bars show 95% bootstrap confidence intervals (1000 subject-level resamples) for balanced logistic regression in each experiment. Across all primary cohorts, severity-only performance lies closer to the reference model than demo-only performance, although the separation between severity-only and demo-only performance is smallest in OASIS3 tau180.

**Fig. 3.**
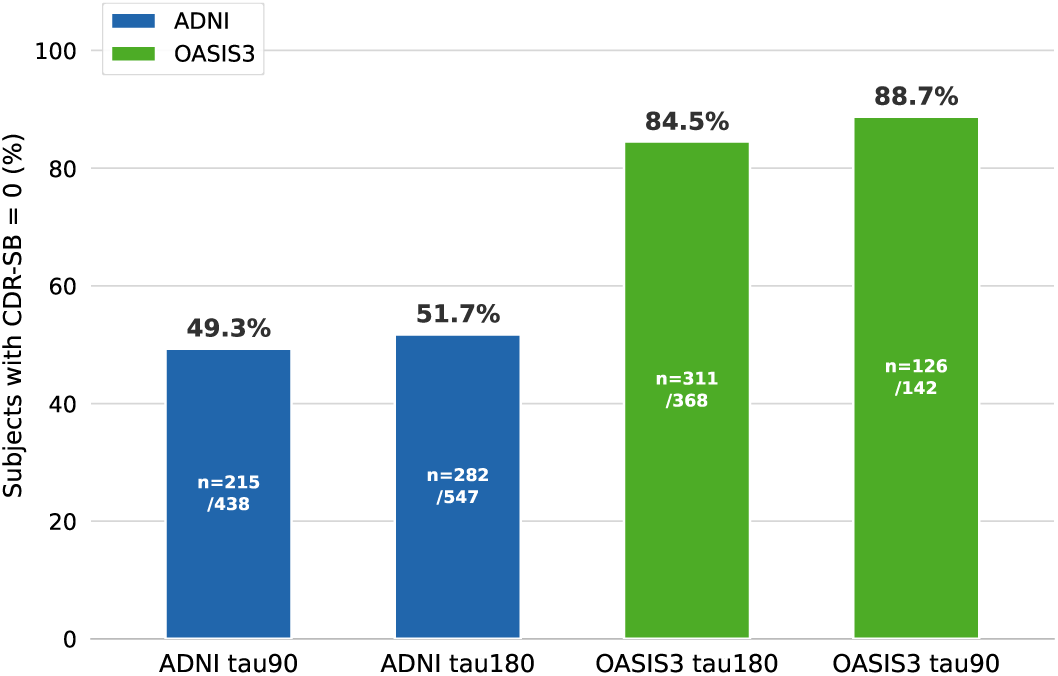
CDR-SB = 0 prevalence by cohort. Bars show the proportion of subjects with CDR-SB = 0, used here as an index of clinically unimpaired enrichment. OASIS3 cohorts were substantially more enriched for functionally intact subjects than ADNI cohorts (84.5–88.7% vs. 49.3–51.7%). The corresponding subgroup counts are shown on each bar. This stage-composition asymmetry provides important context for interpreting why age and APOE retained more independent discriminative value in OASIS3 than in ADNI.

**Table 3.** Primary matched-feature cross-cohort results using balanced logistic regression. Retention fractions express experiment AUC as a proportion of reference AUC. Drop values are reference AUC minus experiment AUC. The matched feature set includes age in all cohorts following age-field repair in ADNI.

| Cohort | $N$ | Pos | AUC | | | Retention | | Drop | |
| --- | --- | --- | --- | --- | --- | --- | --- | --- | --- |
|  |  |  | Ref | Sev | Demo | Sev | Demo | CDR | Full |
| ADNI tau90 | 438 | 78 | 0.851 | 0.838 | 0.743 | 0.984 | 0.873 | 0.094 | 0.108 |
| ADNI tau180 | 547 | 84 | 0.860 | 0.855 | 0.741 | 0.994 | 0.862 | 0.096 | 0.119 |
| OASIS3 tau180 | 368 | 45 | 0.933 | 0.799 | 0.793 | 0.856 | 0.850 | 0.069 | 0.140 |

Ref = reference-combined; Sev = severity-only; Demo = demo-only; CDR drop = $AUC_{ref} - AUC_{minus-all-CDR}$ ; Full drop = $AUC_{ref} - AUC_{demo}$ . Random-forest and extra-trees results are given in Additional file 1.

**Table 4.** Supplementary results. OASIS3 tau90 is small (*n*_pos_ = 12) and should be interpreted cautiously. NACC uses a different endpoint family (strict A+/T+ vs. A*−*/T*−*) and a richer feature set that includes race, Hispanic ethnicity, handedness, category fluency and tau tracer (Section 2.2).

| Cohort | N | Pos | AUC |  |  | Retention |  | Drop |  |
| --- | --- | --- | --- | --- | --- | --- | --- | --- | --- |
|  |  |  | Ref | Sev | Demo | Sev | Demo | CDR | Full |
| OASIS3 tau90 | 142 | 12 | 0.936 | 0.853 | 0.731 | 0.911 | 0.781 | 0.090 | 0.205 |
| NACC strict A/T | 413 | 109 | 0.900 | 0.890 | 0.778 | 0.989 | 0.864 | 0.006 | 0.122 |

Column definitions as in Table 3.

Severity-only models retained most reference signal in all three primary cohorts, with retention fractions of 0.984 (ADNI tau90), 0.994 (ADNI tau180), and 0.856 (OASIS3 tau180). By contrast, demographic/genetic-only models retained less: 0.873, 0.862, and 0.850, respectively. The full-strip drop (reference minus demo-only AUC) was 0.108 (ADNI tau90), 0.119 (ADNI tau180), and 0.140 (OASIS3 tau180).

Bootstrap analysis confirmed that the reference-minus-demo contrast remained positive in all three primary cohorts: 0.108 (95% CI 0.063–0.162) in ADNI tau90, 0.119 (95% CI 0.066–0.168) in ADNI tau180, and 0.140 (95% CI 0.056–0.198) in OASIS3 tau180. By contrast, the reference-minus-severity contrast had confidence intervals crossing zero in both ADNI cohorts: 0.013 (95% CI *−*0.017 to 0.044) in ADNI tau90 and 0.005 (95% CI *−*0.019 to 0.041) in ADNI tau180, indicating that demographic/genetic features added only marginally beyond severity in ADNI under the matched-feature set including age. In OASIS3 tau180 the reference-minus-severity contrast was 0.134 (95% CI 0.058–0.188), indicating that demographic/genetic features add substantial signal beyond severity in the earlier-stage OASIS3 sample.

Removing the CDR-SB block from the reference model reduced performance in ADNI (drops of 0.094 for tau90 and 0.096 for tau180) and in OASIS3 tau180 (drop of 0.069). Removing FAQ alone had minimal effect (drops *<*0.01), indicating that FAQ carried little unique information beyond what CDR-SB and the demographic/genetic features already captured.

### 3.6 Severity retention difference between ADNI and OASIS3

While reference performance differed across cohorts, the severity retention fraction also differed notably: ADNI showed severity retention of 0.984–0.994, whereas OASIS3 tau180 showed 0.856. Under the matched-feature analysis including age, the incremental value of adding severity to the demographic block was 0.108 (95% CI 0.063–0.162) in ADNI tau90 and 0.119 (95% CI 0.066–0.168) in ADNI tau180. Confidence intervals overlapped with OASIS3 tau180 (0.140, 95% CI 0.056–0.198), indicating that the cohort-level difference in severity increment is not statistically distinguishable across all pairwise comparisons.

Notably, the reference-minus-severity contrast had confidence intervals crossing zero in both ADNI cohorts, indicating that demographic/genetic features added only marginally beyond severity in ADNI once age was included in the demographic block. This is distinct from, and should not be confused with, the observation that severity-only AUC exceeds demo-only AUC in ADNI (0.838–0.855 vs. 0.741–0.743): the isolated-block comparison shows that severity alone outperforms demographics alone, whereas the bootstrap contrasts ask what each block contributes uniquely in the combined model. The complementary reference-minus-demo contrast showed that severity added clear information beyond demographics in ADNI, while the reference-minus-severity contrast showed that demographic/genetic features provided little unique information beyond what CDR-SB and FAQ already encoded.

This pattern likely reflects the earlier age-field repair: analyses that excluded age from the demographic block understated demographic/genetic signal and made cross-cohort comparisons sensitive to variable availability. With age properly included, the ADNI results show an asymmetric pattern: severity adds clear information beyond demographics, whereas demographics add little beyond severity.

These patterns are consistent with the disease-stage composition asymmetry documented in Section 3.3: in OASIS3, where 84.5% of subjects had CDR-SB = 0, severity features alone are insufficient to discriminate tau positivity for the majority who have no measurable impairment, making the demographic block—particularly age and APOE—essential for prediction. In ADNI, where roughly half the sample has measurable functional impairment, severity already encodes most of the discriminative information and the demographic block adds little beyond it. The higher reference AUC in OASIS3 reflects the stronger joint discriminative power of the full feature set in an earlier-stage sample, not a fundamental difference in biology.

### 3.7 Diagnosis-stratified feature-block analysis

To directly test whether the severity-entanglement pattern varied by disease stage, we repeated the reference-combined and demo-only experiments within diagnosis strata defined by baseline clinical classification. Strata with fewer than 10 tau-positive or 10 tau-negative subjects were not estimated; this excluded the ADNI CN strata in both primary cohorts (*N*_+_ = 6) and the OASIS3 MCI stratum (*N*_+_ = 4). Diagnosis distributions were: ADNI tau90 CN *n* = 235, MCI *n* = 151, Dementia *n* = 52; ADNI tau180 CN *n* = 312, MCI *n* = 176, Dementia *n* = 59; OASIS3 tau180 CN *n* = 318, MCI *n* = 9, Dementia *n* = 37 (four OASIS3 subjects with other or missing diagnosis were excluded from stratified analyses).

In OASIS3 tau180, the pattern reversed by stratum in the expected direction. In the CN stratum (*N*_+_ = 16), demo-only AUC (0.841) exceeded reference AUC (0.799), suggesting that severity features added noise rather than signal when nearly all subjects had CDR-SB = 0. In the Dementia stratum (*N*_+_ = 25, *N_−_* = 12), reference AUC (0.660) exceeded demo-only AUC (0.493), consistent with severity features providing discriminative information when functional impairment was present, although with only 12 tau-negative subjects these Dementia-stratum estimates are imprecise.

In ADNI, Dementia strata were estimable (*N*_+_ = 36–39 and *N_−_* = 16–20 across tau90/tau180). In ADNI tau90 Dementia, reference AUC (0.839) modestly exceeded demo-only (0.827), consistent with severity providing incremental information. In ADNI tau180 Dementia, the pattern was directionally inconsistent (reference 0.733 vs. demo-only 0.786), suggesting the severity advantage in the Dementia stratum is weaker and less consistent across ADNI cohorts than the stratum reversal observed in OASIS3. ADNI MCI strata (*N*_+_ = 36–39) showed demo-only AUC at or above reference (demo 0.662–0.688 vs. reference 0.647–0.662), suggesting that the severity block provides limited additional signal in intermediate-stage subjects.

The OASIS3 CN-to-Dementia reversal is consistent with the signature predicted by severity entanglement: in a stratum where CDR-SB is near-uniformly zero, adding severity features to the demographic block introduces noise rather than signal, reducing AUC below the demo-only level. In the Dementia stratum, where functional impairment varies substantially, severity provides discriminative information that demographics alone cannot capture. These stratum-level results are presented in Supplementary Fig. S2.

### 3.8 Supplementary cohorts

OASIS3 tau90 showed the same directional pattern but was considered unstable due to only 12 positive cases. In this cohort, reference AUC was 0.936, severity-only was 0.853, and demo-only was 0.731 (Table 4).

NACC strict A/T served as an endpoint-family sensitivity anchor. Because this endpoint family and feature set differ from the matched ADNI/OASIS3 tau-positivity analysis, NACC was used as a sensitivity anchor rather than as a directly pooled primary cohort. With a different endpoint (strict A+/T+ vs. A*−*/T*−*), a richer feature set (including CDR global, MoCA, category fluency, race, Hispanic ethnicity, handedness, and tau tracer), and higher positivity rate (26.4%), NACC showed reference AUC of 0.900, severity-only of 0.890, and demo-only of 0.778. Part of the demo-only signal reflected tracer identity, because A+/T+ prevalence differed by tracer (32.8% with flortaucipir vs. 10.3% with MK-6240): without the tracer term, demo-only AUC was 0.747, and with only the five harmonized ADNI/OASIS3 demographic/genetic variables it was 0.712 (Additional file 4). The qualitative pattern of severity retaining more reference signal than demographics alone was preserved, though the NACC end-point family, richer feature set, and higher positivity rate preclude direct comparison with the primary ADNI–OASIS3 analysis. Notably, the CDR block drop in NACC was very small (0.006), suggesting that the remaining severity measures (MoCA, FAQ, and category fluency) largely substituted for CDR; removing CDR, FAQ, and MoCA together reduced AUC by 0.119.

## 4 Discussion

### 4.1 Summary of findings

This study was designed as a methodological investigation rather than a prediction benchmark. The results follow a clear two-part sequence. First, the original large ADNI–OASIS3 performance gap was largely a feature-availability artifact: the pre-harmonization demo-only gap of 0.204 (ADNI tau180 0.742 vs. OASIS3 0.538) arose because OASIS3 lacked age and APOE in its demographic block. After feature audit, age-field repair, and matched-feature restriction, the absolute gap narrowed to 0.052 and the signed ADNI-minus-OASIS3 difference reversed direction (ADNI 0.741 vs. OASIS3 0.793). Second, the residual reversed gap after harmonization is most consistent with cohort disease-stage composition asymmetry: the OASIS3 sample was predominantly clinically unimpaired (84.5% CDR-SB = 0 in tau180), where age and APOE carry stronger discriminative power because severity features are largely uninformative for the majority of subjects, whereas the ADNI sample was substantially more severity-enriched (49–52% CDR-SB = 0). This two-part interpretation is more precise than attributing the gap to either feature availability or stage structure alone, and it supports the broader conclusion that naive cross-cohort comparisons can substantially overstate biological differences when cohorts differ in both variable availability and disease-stage composition.

### 4.2 Severity as a major but stage-dependent predictor

Clinical severity remained an important contributor to tau-positivity prediction in every cohort examined. Severity-only performance was close to the reference-combined model across all primary analyses, with retention fractions ranging from 0.856 to 0.994. Demographic/genetic-only performance was consistently lower, with retention fractions of 0.850 to 0.873. However, the incremental value of demographics above severity was modest and statistically uncertain in both ADNI cohorts once age was properly included in the matched feature set, whereas the corresponding increment was larger in OASIS3. Thus, severity remained a major predictor, but the degree to which severity dominated over demographic/genetic information depended strongly on disease-stage composition.

This finding has a dual interpretation. On one hand, it is unsurprising: tau pathology drives clinical severity, so clinical severity measures should correlate with tau positivity. On the other hand, it raises a concern about what tau-prediction models are actually learning. Models that rely heavily on CDR-SB, FAQ, or similar measures may be predicting downstream clinical consequences of pathology as much as identifying upstream biological risk. This distinction matters when such models are used to assess the value of demographic or genetic predictors, because residual demographic signal (after severity is accounted for) is more biologically informative than raw demographic signal that is confounded with severity.

Logistic regression coefficients from the primary matched-feature reference model showed that CDR-SB had the largest coefficient in every primary cohort (standardized coefficient 1.37–1.69). In ADNI it was more than twice the largest demographic/genetic coefficient (APOE *ε*4 count, 0.49–0.59); in OASIS3 tau180, age (1.33) and APOE *ε*4 count (1.07) approached CDR-SB (1.69), consistent with the earlier-stage composition of that cohort. FAQ carried a negative coefficient after conditioning on CDR-SB in all three primary cohorts, consistent with collinearity between the two severity measures. The female-versus-male contrast also differed between cohorts (1.02 in OASIS3 vs. *−*0.15 to *−*0.18 in ADNI); with 45 tau-positive OASIS3 participants we do not interpret this difference further (Supplementary Table S6).

An important caveat in interpreting cross-cohort severity differences concerns the clinical meaning of CDR-SB = 0. A CDR-SB score of zero indicates no detectable functional impairment across any rated domain; it does not establish that a subject is cognitively normal. Most CDR-SB = 0 subjects in our cohorts were tau-negative (tau-positive prevalence 1.8–4.8% in this stratum; Section 3.4); only the small tau-positive subset corresponds to preclinical or very early prodromal AD. We therefore use CDR-SB = 0 as an index of clinically unimpaired enrichment rather than of preclinical disease: approximately half of ADNI subjects met this definition (49–52%), compared with 84.5–88.7% of OASIS3 subjects.

This stage asymmetry has two distinct but related implications, which should not be conflated. First, severity-only AUC exceeded demo-only AUC in both ADNI cohorts (0.838–0.855 vs. 0.741–0.743), meaning that severity features alone outperform demographics alone in ADNI; this reflects the high prevalence of measurable functional impairment in a biomarker-enriched cohort, where CDR-SB carries strong discriminative information. Second, and separately, the reference-minus-severity contrast shows that once both blocks are included, demographics added only marginally beyond severity in ADNI (contrasts 0.013 and 0.005, both with CIs crossing zero), while demographics added substantial signal beyond severity in OASIS3 (0.134, 95% CI 0.058–0.188). The first comparison concerns which block is more informative in isolation; the second concerns what each block contributes uniquely in a combined model. The severity-only AUC difference between ADNI tau180 and OASIS3 tau180 (0.855 vs. 0.799) is consistent with the disease-stage composition asymmetry rather than a fundamental difference in how well clinical severity tracks tau burden. Our CDR-SB = 0 sensitivity analysis supported this interpretation by showing a marked collapse in tau-positive prevalence within the functionally intact subgroup, leaving too few ADNI tau-positive cases for reliable within-stage model estimation. Future work with larger samples should repeat this comparison within adequately powered stage-matched strata or using continuous tau burden.

The diagnosis-stratified analysis (Section 3.7) provided supportive evidence for this mechanism: in the OASIS3 CN stratum, where CDR-SB was near-uniformly zero, adding severity features to the demographic block reduced AUC rather than improving it (demo-only 0.841 vs. reference 0.799), whereas in the Dementia stratum severity features added discriminative value (reference 0.660 vs. demo-only 0.493), although the corresponding ADNI Dementia pattern was weaker and directionally inconsistent. This stratum reversal matches the pattern predicted by severity entanglement, but the small OASIS3 Dementia stratum (*N_−_*= 12) limits how much weight it can bear.

A related finding from the matched-feature rerun is that including age clarified the direction of unique contributions in the combined model. The reference-minus-severity contrast in both ADNI cohorts had confidence intervals crossing zero (0.013, 95% CI *−*0.017 to 0.044 in ADNI tau90; 0.005, 95% CI *−*0.019 to 0.041 in ADNI tau180), indicating that once age is a legitimate covariate, demographics add only marginally beyond CDR-SB and FAQ in ADNI. In OASIS3 tau180, by contrast, the same contrast was 0.134 (95% CI 0.058–0.188), indicating that demographic/genetic features add substantial signal beyond severity in the earlier-stage OASIS3 sample. This asymmetry is consistent with disease-stage composition differences: in a predominantly clinically unimpaired sample, age and APOE retain discriminative power that is already largely captured by severity measures in a more severity-enriched cohort.

### 4.3 Relationship to tau–clinical mismatch literature

Our finding that severity-only models capture 86–99% of reference performance is broadly consistent with the tau–clinical concordance observed in recent mismatch analyses. Brown et al. found that approximately 56% of amyloid-positive individuals in ADNI showed canonical concordance between tau burden and clinical severity, while 24% were classified as resilient and 20% as vulnerable [13]. Our ablation results are broadly consistent with this picture: severity captures most but not all of the tau-positivity signal, and the residual gap (1–14% of reference AUC, depending on cohort) may partly correspond to signal from the non-canonical subpopulations, though direct attribution requires analyses not conducted here.

### 4.4 Harmonization as a scientific determinant

The feature audit and age-field repair provided an important practical lesson. Before variable recovery, OASIS3 appeared to have substantially weaker demographic-only prediction than ADNI (AUC 0.538 vs. 0.742). After recovering APOE and age in OASIS3, and repairing the age field in ADNI, the absolute demo-only AUC gap shrank from 0.204 to 0.052 and the signed ADNI-minus-OASIS3 difference reversed direction. This is most consistent with the interpretation that the original gap was largely a feature-availability artifact, with the residual difference after harmonization likely reflecting disease-stage composition asymmetry—though the contribution of endpoint-definition differences cannot be excluded. Researchers reporting cross-cohort tau prediction results should explicitly document which variables were available in each cohort, whether apparently missing variables were audited for recoverability, and whether numeric fields were validated for plausibility before use.

### 4.5 Limitations

This work has several limitations.

First, the OASIS3 tau90 cohort contained only 12 positive cases, yielding approximately 2–3 positives per cross-validation fold. AUC estimates at this sample size carry wide confidence intervals, and this cohort was therefore retained only as supplementary.

Second, the matched-feature analysis was constrained by the intersection of recoverable variables. MoCA and CDR global were absent from the OASIS3 extract. A richer shared feature set would provide a more complete picture of cross-cohort signal structure.

Third, although we estimated bootstrap confidence intervals for the primary AUC contrasts, these intervals remain fairly wide, especially in OASIS3 tau180. Accordingly, the cross-cohort AUC comparisons should be interpreted as directional rather than precise.

Fourth, tau-positivity definitions differed between cohorts. ADNI used a pre-existing label derived from established flortaucipir SUVR thresholds, whereas OASIS3 used a data-driven GMM midpoint threshold on the harmonized tauopathy SUVR summary variable. We did not adopt the published OASIS-3 cutoff (1.22) as the primary definition because in our cohorts it labelled nearly half of participants as tau-positive, about half of whom were amyloid-negative (Section 2.2; Supplementary Table S5). Although threshold-sensitivity analysis suggested that OASIS3 tau180 demographic discriminability was not driven by a single brittle cut point, endpoint-definition asymmetry remains an important limitation and may contribute to the higher reference AUC observed in OASIS3. A within-OASIS3 continuous-SUVR sensitivity analysis showed that the severity signal persisted without binary thresholding, but this does not eliminate cross-cohort endpoint asymmetry because continuous ADNI and OASIS3 SUVR values are not on a common scale across tracers, preprocessing pipelines, and reference regions. Future work should evaluate continuous and regional tau burden using formally harmonized PET measures or tracer-specific calibration models. A common endpoint, such as visual reads applied with the same criteria in ADNI and OASIS-3 [17], would remove this asymmetry and would also make cross-cohort transfer experiments interpretable.

Fifth, the study focused on tabular clinical and genetic predictors. Imaging features (MRI volumetrics, PET SUVRs) and plasma biomarkers were not included because the present objective was interpretability of cross-cohort signal structure using widely available clinical variables.

Sixth, we did not perform cross-cohort transfer experiments (training on one cohort and testing on another). While such experiments would be informative in principle, the sample sizes—particularly in OASIS3—and the endpoint-definition asymmetry would make transfer results difficult to interpret cleanly.

Seventh, all reported AUCs are cross-validated estimates from five-fold stratified cross-validation; no separate held-out test set was withheld, as sample sizes (*N*_+_ = 45–84 per primary cohort) would render held-out evaluation underpowered. Cross-validated AUC may be modestly optimistic relative to true generalization performance, and results should be interpreted accordingly.

### 4.6 Future directions

The most important next step is to extend this framework to formally harmonized continuous and regional tau measures across cohorts rather than relying on binary positivity. The within-OASIS3 continuous-SUVR sensitivity analysis reported here suggests that severity-stage structure persists without binary thresholding, but cross-cohort continuous tau comparisons will require tracer-aware harmonization and common reference-region calibration. Binary thresholding discards substantial information about tau severity and spatial distribution, and the field is moving toward continuous and regional characterization [3, 4]. A second direction is adequately powered stage-stratified or stage-matched analysis, particularly within larger CDR-SB = 0 samples or using continuous tau burden. A third direction is to incorporate plasma biomarkers (p-tau217, NfL, GFAP) into the ablation framework, as these represent a biologically upstream predictor domain that is less susceptible to severity entanglement.

## 5 Conclusion

Matched-feature cross-cohort analysis showed that the apparent ADNI–OASIS3 gap in tau-positivity prediction followed a two-part pattern. The original large gap was largely a feature-availability artifact, with the absolute gap narrowing and the signed ADNI-minus-OASIS3 difference reversing after feature audit, age-field repair, and matched-feature restriction. The residual reversed difference after harmonization was most consistent with cohort disease-stage composition asymmetry: OASIS3 was predominantly clinically unimpaired (84.5% CDR-SB = 0 in tau180), where age and APOE carry stronger discriminative power, while ADNI was substantially more severity-enriched, where severity already captures most of the predictive signal and demographics add little beyond it. Diagnosis-stratified analyses were consistent with this severity-entanglement mechanism: in the OASIS3 CN stratum, severity features reduced rather than improved performance over demographics alone, whereas in the Dementia stratum they added discriminative information; the corresponding ADNI Dementia pattern was weaker and directionally inconsistent across matching windows. These findings argue that cross-cohort biomarker modeling should account for both feature-set completeness and disease-stage composition heterogeneity, not only headline performance metrics.

## Data Availability

Data used in this study are available to qualified investigators from ADNI (https://adni.loni.usc.edu/), OASIS (https://www.oasis-brains.org/) and NACC (https://naccdata.org/) under their data use agreements. Analysis code is available at https://github.com/abhatfnal/crosscohort-tau-harmonization.

https://www.oasis-brains.org/

https://naccdata.org/

https://github.com/abhatfnal/crosscohort-tau-harmonization

## Supplementary information

Additional file 1: Full matched-feature rerun metrics across all experiments and model families. Additional file 2: OASIS feature-audit tables and recovered column mappings. Additional file 3: Full-feature (unmatched) ADNI feature-block ablation results. Additional file 4: NACC sensitivity results (balanced logistic regression), including a flortaucipir-only subset and demographic-block variants without tau tracer. Additional file 5: OASIS3 GMM threshold-sensitivity results, including Supplementary Fig. S1 and Supplementary Table S5. Additional file 6: Standardized logistic regression feature importance coefficients across cohorts and experiments (Supplementary Table S6). Additional file 7: Diagnosis-stratified feature-block AUC results across CN, MCI, and Dementia strata (Supplementary Fig. S2).

Additional file 8: Within-OASIS3 continuous tauopathy SUVR sensitivity analysis, including Supplementary Table S7 and Supplementary Fig. S3.

## Acknowledgements

Data collection and sharing for the Alzheimer’s Disease Neuroimaging Initiative (ADNI) is funded by the National Institute on Aging (National Institutes of Health Grant U19AG024904). The grantee organization is the Northern California Institute for Research and Education. In the past, ADNI has also received funding from the National Institute of Biomedical Imaging and Bioengineering, the Canadian Institutes of Health Research, and private sector contributions through the Foundation for the National Institutes of Health (FNIH), including AbbVie, Alzheimer’s Association, Alzheimer’s Drug Discovery Foundation, Araclon Biotech, BioClinica, Inc., Biogen, Bristol-Myers Squibb Company, CereSpir, Inc., Cogstate, Eisai Inc., Elan Pharmaceuticals, Inc., Eli Lilly and Company, EuroImmun, F. Hoffmann-La Roche Ltd and its affiliated company Genentech, Inc., Fujirebio, GE Healthcare, IXICO Ltd., Janssen Alzheimer Immunotherapy Research & Development, LLC, Johnson & Johnson Pharmaceutical Research & Development LLC, Lumosity, Lundbeck, Merck & Co., Inc., Meso Scale Diagnostics, LLC, NeuroRx Research, Neurotrack Technologies, Novartis Pharmaceuticals Corporation, Pfizer Inc., Piramal Imaging, Servier, Takeda Pharmaceutical Company, and Transition Therapeutics.

Data were provided in part by OASIS-3: Longitudinal Multimodal Neuroimaging. Principal Investigators: T. Benzinger, D. Marcus, and J. Morris; supported by NIH grants P30 AG066444, P50 AG00561, P30 NS09857781, P01 AG026276, P01 AG003991, R01 AG043434, UL1 TR000448, and R01 EB009352. AV-45 doses were provided by Avid Radiopharmaceuticals, a wholly owned subsidiary of Eli Lilly.

OASIS-3 AV1451 data were provided in part by OASIS-3 AV1451: Principal Investigators T. Benzinger and J. Morris; supported by NIH P30 AG066444 and AW00006993. AV-1451 doses were provided by Avid Radiopharmaceuticals, a wholly owned subsidiary of Eli Lilly.

The NACC database is funded by NIA/NIH Grant U24 AG072122. NACC data are contributed by the NIA-funded ADRCs: P30 AG062429 (PI James Brewer, MD, PhD), P30 AG066468 (PI Oscar Lopez, MD), P30 AG062421 (PI Bradley Hyman, MD, PhD), P30 AG066509 (PI Thomas Grabowski, MD), P30 AG066514 (PI Mary Sano, PhD), P30 AG066530 (PI Helena Chui, MD), P30 AG066507 (PI Marilyn Albert, PhD), P30 AG066444 (PI David Holtzman, MD), P30 AG066518 (PI Lisa Silbert, MD, MCR), P30 AG066512 (PI Thomas Wisniewski, MD), P30 AG066462 (PI Scott Small, MD), P30 AG072979 (PI David Wolk, MD), P30 AG072972 (PI Charles DeCarli, MD), P30 AG072976 (PI Andrew Saykin, PsyD), P30 AG072975 (PI Julie A. Schneider, MD, MS), P30 AG072978 (PI Ann McKee, MD), P30 AG072977 (PI Robert Vassar, PhD), P30 AG066519 (PI Frank LaFerla, PhD), P30 AG062677 (PI Ronald Petersen, MD, PhD), P30 AG079280 (PI Jessica Langbaum, PhD), P30 AG062422 (PI Gil Rabinovici, MD), P30 AG066511 (PI Allan Levey, MD, PhD), P30 AG072946 (PI Linda Van Eldik, PhD), P30 AG062715 (PI Sanjay Asthana, MD, FRCP), P30 AG072973 (PI Russell Swerdlow, MD), P30 AG066506 (PI Glenn Smith, PhD, ABPP), P30 AG066508 (PI Stephen Strittmatter, MD, PhD), P30 AG066515 (PI Victor Henderson, MD, MS), P30 AG072947 (PI Suzanne Craft, PhD), P30 AG072931 (PI Henry Paulson, MD, PhD), P30 AG066546 (PI Sudha Seshadri, MD), P30 AG086401 (PI Erik Roberson, MD, PhD), P30 AG086404 (PI Gary Rosenberg, MD), P20 AG068082 (PI Angela Jefferson, PhD), P30 AG072958 (PI Heather Whitson, MD), P30 AG072959 (PI James Leverenz, MD).

We thank institutional computing support at the University of Chicago Research Computing Center.

## Abbreviations

AD: Alzheimer’s disease
ADNI: Alzheimer’s Disease Neuroimaging Initiative
ADRC: Alzheimer’s Disease Research Center
AP: average precision
APOE: apolipoprotein E
AUC: area under the receiver operating characteristic curve
CDR-SB: CDR^®^ Dementia Staging Instrument sum of boxes
CI: confidence interval
CN: cognitively normal
CV: cross-validation
FAQ: Functional Activities Questionnaire
GFAP: glial fibrillary acidic protein
GMM: Gaussian mixture model
MCI: mild cognitive impairment
MoCA: Montreal Cognitive Assessment
MRI: magnetic resonance imaging
NACC: National Alzheimer’s Coordinating Center
NfL: neurofilament light chain
OASIS3: Open Access Series of Imaging Studies 3
PET: positron emission tomography
p-tau217: tau phosphorylated at threonine 217
SCAN: Standardized Centralized Alzheimer’s and Related Dementias Neuroimaging
SD: standard deviation
SUVR: standardized uptake value ratio

## Declarations

### Ethics approval and consent to participate

This study is a secondary analysis of de-identified data obtained under the data use agreements of ADNI, OASIS-3, and NACC; the authors had no interaction with participants and no access to identifying information. ADNI was approved by the institutional review boards of all participating sites, and written informed consent was obtained from all participants or their authorized representatives. OASIS-3 data collection was approved by the Washington University in St. Louis Institutional Review Board, and all participants provided informed consent. NACC data are collected under protocols approved by the institutional review board of each contributing Alzheimer’s Disease Research Center, with informed consent from all participants.

### Consent for publication

Not applicable.

### Availability of data and materials

ADNI data are available from the Alzheimer’s Disease Neuroimaging Initiative repository (https://adni.loni.usc.edu/) to qualified investigators who complete the required data use agreement. OASIS-3 data are available from OASIS (https://www.oasis-brains.org/) to qualified investigators under the applicable data use terms. NACC data are available from the National Alzheimer’s Coordinating Center (https://naccdata.org/) to qualified investigators under the applicable data request and data use procedures. Individual participant-level data are not redistributed by the authors. Analysis code used for harmonization, feature audit, matched-feature reruns, bootstrap confidence-interval estimation, and figure generation is available at https://github.com/abhatfnal/crosscohort-tau-harmonization. Code and selected rendered aggregate outputs supporting the manuscript figures and tables are shared; no participant-level ADNI, OASIS, or NACC data, and no controlled-access participant-derived intermediate tables, are publicly redistributed.

### Authors’ contributions

A. B. conceived the study, performed all analyses, and drafted the manuscript. M.C. contributed to study design and interpretation. C.-T.C. contributed to study design and scientific interpretation. B.T.F. and G.T.F. supervised the work and revised the manuscript. All authors read and approved the final manuscript.

---

**Fig. S1.**
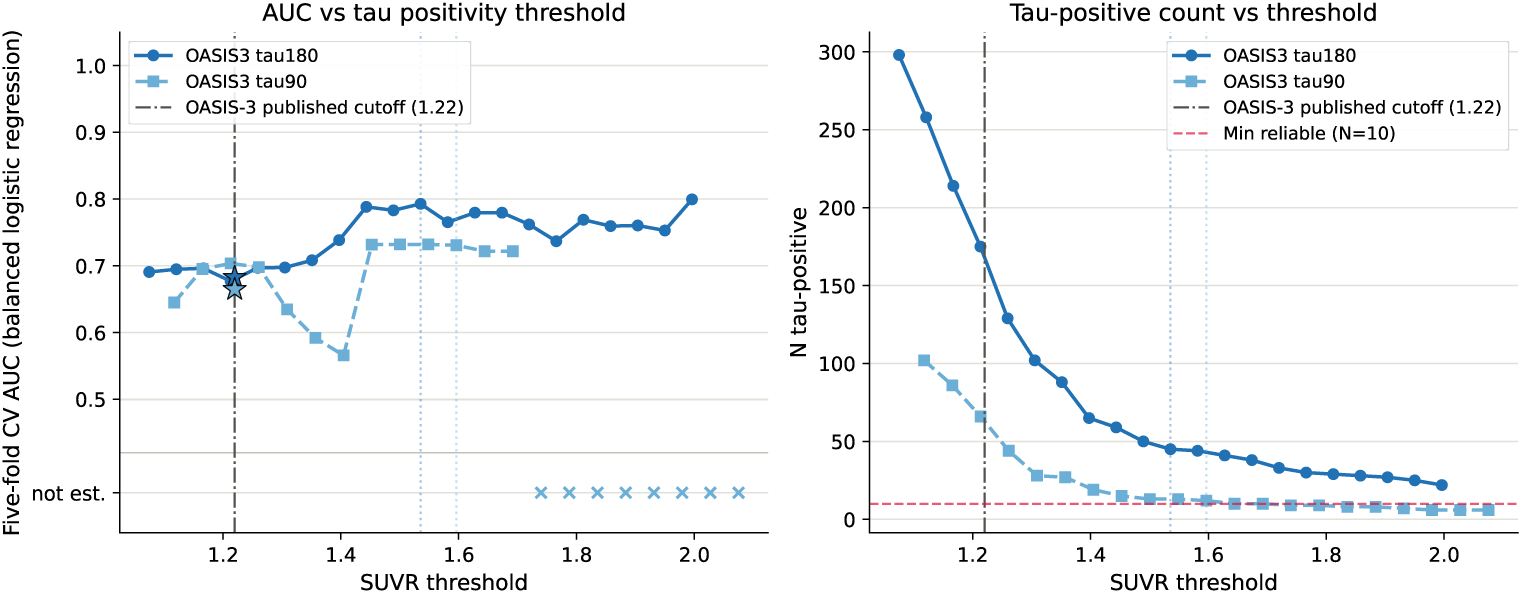
OASIS3 tau-threshold sensitivity analysis. Threshold-sensitivity analysis for OASIS3 tau positivity using demographic/genetic predictors only: age, APOE *ε*4 count, APOE *ε*4 carrier status, education, and sex. Left, AUC versus threshold; right, number of tau-positive subjects versus threshold. Vertical dotted lines mark the cohort-specific Gaussian mixture model midpoint thresholds used to derive tau positivity; the dash-dotted line and stars mark the published OASIS-3 cutoff (1.22). Points indicate mean five-fold cross-validated AUC from balanced logistic regression at each threshold. Thresholds yielding fewer than 10 tau-positive subjects (red dashed line, right) are marked *×* in the ‘not est.’ band. In OASIS3 tau180, demographic discriminability remained directionally stable near the GMM midpoint, where the tau-positive count matched the primary cohort definition (*n* = 45). In OASIS3 tau90, the GMM midpoint yielded 12 tau-positive subjects and a demographic/genetic AUC of 0.731, but the cohort remained too small for primary inference.

**Fig. S2.**
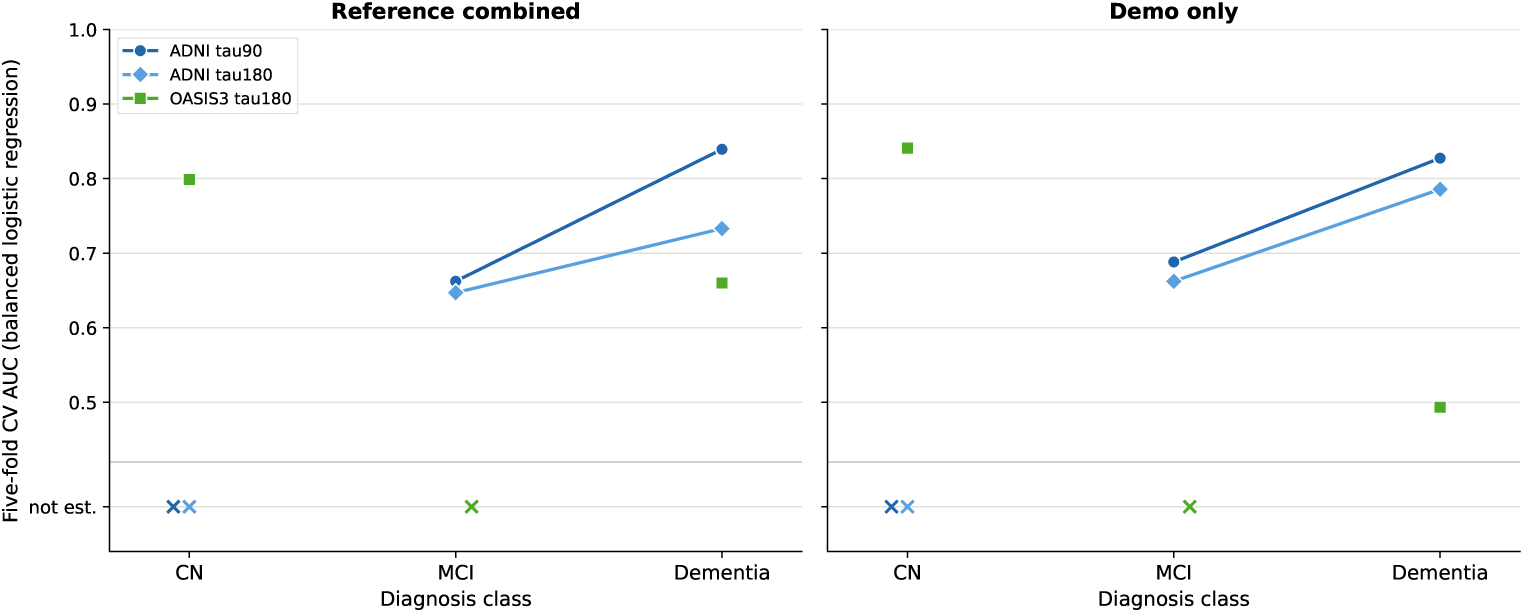
Diagnosis-stratified feature-block AUC analysis. Reference-combined and demo-only AUCs stratified by baseline diagnosis (CN, MCI, Dementia) for ADNI tau90, ADNI tau180, and OASIS3 tau180 (balanced logistic regression, five-fold cross-validation). Strata with fewer than 10 tau-positive or 10 tau-negative subjects were not estimated and are marked *×* in the ‘not est.’ band (ADNI CN: *N*_+_ = 6; OASIS3 MCI: *N*_+_ = 4). In OASIS3 tau180, the CN stratum shows demo-only AUC (0.841) exceeding reference AUC (0.799), suggesting that severity features add noise when CDR-SB is near-uniformly zero; the Dementia stratum shows the pattern reversing (reference 0.660 vs. demo-only 0.493; *N*_+_ = 25, *N_−_* = 12), consistent with severity contributing discriminative information when functional impairment is present. In ADNI, the severity advantage in the Dementia stratum was present in tau90 (reference 0.839 vs. demo-only 0.827) but directionally inconsistent in tau180 (reference 0.733 vs. demo-only 0.786), indicating a weaker and less consistent effect than in OASIS3.

**Fig. S3.**
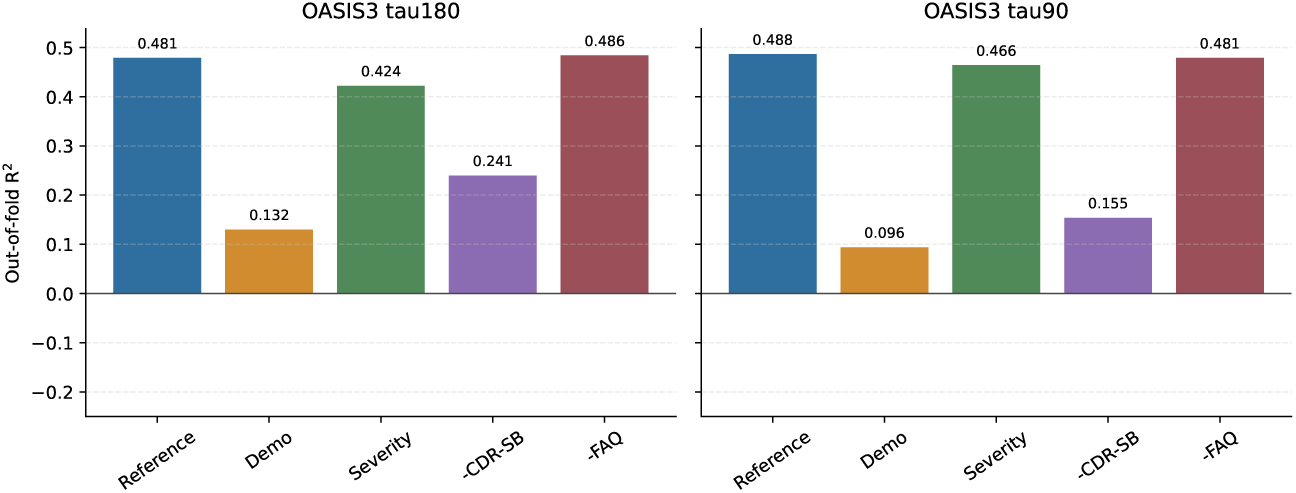
Within-OASIS3 continuous tauopathy SUVR sensitivity analysis. Cross-validated *R*^2^ for ridge-regression models predicting continuous Tauopathy SUVR from matched feature blocks in OASIS3 tau90 and tau180. The reference-combined and severity-only models explained substantially more variance than the demo-only model, indicating that the OASIS3 severity-stage signal persisted when tauopathy was modeled as a continuous outcome rather than binarized at the GMM midpoint.

**Table S5.** Selected OASIS3 threshold-sweep results around the GMM midpoint using the harmonized tauopathy SUVR variable. AUCs are from demographic/genetic-only balanced logistic regression using age, APOE *ε*4 count, APOE *ε*4 carrier status, education, and sex. The first row of each block uses the published OASIS-3 tauopathy cutoff (1.22) [16]. Reliable thresholds are defined as those with at least 10 tau-positive subjects. AUCs use a single five-fold split (seed 42), as in the primary analysis; across 100 split seeds the standard deviation at the thresholds checked was about 0.01 (tau180) and 0.02 (tau90).

| Cohort | % from GMM | Threshold | $N$ | Tau+ | AUC | Reliable |
| --- | --- | --- | --- | --- | --- | --- |
| OASIS3 tau180 | −20.6% (published) | 1.220 | 368 | 171 | 0.683 | Yes |
| OASIS3 tau180 | −9% | 1.397 | 368 | 65 | 0.738 | Yes |
| OASIS3 tau180 | −6% | 1.443 | 368 | 59 | 0.788 | Yes |
| OASIS3 tau180 | −3% | 1.489 | 368 | 50 | 0.783 | Yes |
| OASIS3 tau180 | 0% | 1.536 | 368 | 45 | 0.793 | Yes |
| OASIS3 tau180 | +3% | 1.582 | 368 | 44 | 0.765 | Yes |
| OASIS3 tau180 | +6% | 1.628 | 368 | 41 | 0.780 | Yes |
| OASIS3 tau90 | −23.6% (published) | 1.220 | 142 | 64 | 0.665 | Yes |
| OASIS3 tau90 | −9% | 1.453 | 142 | 15 | 0.732 | Yes |
| OASIS3 tau90 | −6% | 1.501 | 142 | 13 | 0.732 | Yes |
| OASIS3 tau90 | −3% | 1.549 | 142 | 13 | 0.732 | Yes |
| OASIS3 tau90 | 0% | 1.596 | 142 | 12 | 0.731 | Yes |
| OASIS3 tau90 | +3% | 1.644 | 142 | 10 | 0.722 | Yes |
| OASIS3 tau90 | +6% | 1.692 | 142 | 10 | 0.722 | Yes |
| OASIS3 tau90 | +9% | 1.740 | 142 | 9 | — | No |

**Table S6.** Standardized logistic regression coefficients from the primary matched-feature reference-combined model, fitted on the full cohort. Numeric features, including the binary APOE *ε*4 carrier indicator, were median-imputed and z-scored, so coefficients are changes in log-odds per one-SD increase; sex is the difference between the female and male one-hot coefficients (log-odds, not per SD). CDR-SB had the largest coefficient in every cohort (1.37–1.69). In ADNI it was more than twice as large as any demographic/genetic coefficient (largest: APOE *ε*4 count, 0.49–0.59); in OASIS3, age (1.33) and APOE *ε*4 count (1.07) approached CDR-SB, consistent with the earlier disease-stage composition of that sample. APOE *ε*4 carrier status and allele count are highly collinear (*r* = 0.92–0.93), so their individual coefficients (including the negative OASIS3 carrier coefficient) should not be interpreted separately. The negative FAQ coefficient after conditioning on CDR-SB likewise reflects collinearity between the two severity measures.

| Feature | ADNI tau90 | ADNI tau180 | OASIS3 tau180 |
| --- | --- | --- | --- |
| CDR-SB | 1.37 | 1.40 | 1.69 |
| Age | 0.23 | 0.15 | 1.33 |
| APOE ε4 carrier | 0.25 | 0.15 | −0.16 |
| APOE ε4 count | 0.49 | 0.59 | 1.07 |
| Education | −0.18 | −0.15 | 0.08 |
| Sex (female vs. male)<sup>a</sup> | −0.18 | −0.15 | 1.02 |
| FAQ | −0.24 | −0.19 | −0.19 |

<sup>a</sup>Difference between the female and male one-hot coefficients (log-odds, not per SD).

**Table S7.** Within-OASIS3 continuous tauopathy SUVR sensitivity analysis. Ridge-regression models were fit using continuous Tauopathy SUVR as the outcome and the same matched feature blocks as in the binary analysis. Performance is reported as five-fold cross-validated *R*^2^. This analysis was performed only within OASIS3 and was not used for direct cross-cohort comparison.

| Cohort | Reference | Demo-only | Severity-only | Minus all CDR* | Minus FAQ |
| --- | --- | --- | --- | --- | --- |
| OASIS3 tau180 | 0.481 | 0.132 | 0.424 | 0.241 | 0.486 |
| OASIS3 tau90 | 0.488 | 0.096 | 0.466 | 0.155 | 0.481 |

\*In the matched OASIS3 feature set, minus-all-CDR is operationalized as removing CDR-SB.

## References

[1] Ossenkoppele, R., Pichet Binette, A., Groot, C., et al.: Amyloid and tau PET-positive cognitively unimpaired individuals are at high risk for future cognitive decline. Nature Medicine 28(11), 2381–2387 (2022) 10.1038/s41591-022-02049-x

[2] Moscoso, A., Heeman, F., Raghavan, S., et al.: Frequency and clinical outcomes associated with tau positron emission tomography positivity. JAMA 334(3), 229–242 (2025) 10.1001/jama.2025.7817

[3] Karlsson, L., Vogel, J.W., Arvidsson, I., et al.: Machine learning prediction of tau-PET in Alzheimer’s disease using plasma, MRI, and clinical data. Alzheimer’s & Dementia 21(2), 14600 (2025) 10.1002/alz.14600

[4] Rathore, S., Habes, M., Palsetia, D., et al.: Predicting regional tau accumulation with machine learning-based tau-PET and advanced radiomics. Alzheimer’s & Dementia: Translational Research & Clinical Interventions 10(4), 70005 (2024) 10.1002/trc2.70005

[5] Ang, T.-F.A., Joshi, P., Katz, M.J., et al.: Development of simple risk scores for prediction of brain *β*-amyloid and tau status in older adults with mild cognitive impairment: a machine learning approach. Alzheimer’s & Dementia: Diagnosis, Assessment & Disease Monitoring 17(1), 70116 (2025) 10.1002/dad2.70116

[6] Lan, G., Wang, Y., Peng, W., et al.: Comprehensive evaluation of plasma tau biomarkers for detecting and monitoring Alzheimer’s disease in a multicenter and multiethnic aging population. Nature Aging 5(8), 1601–1618 (2025) 10.1038/s43587-025-00904-3

[7] Bourgeat, P., Doré, V., Burnham, S.C., et al.: *β*-amyloid PET harmonisation across longitudinal studies: application to AIBL, ADNI and OASIS3. NeuroImage 262, 119585 (2022) 10.1016/j.neuroimage.2022.119585

[8] Landau, S.M., Harrison, T.M., Baker, S.L., et al.: Positron emission tomography harmonization in the Alzheimer’s Disease Neuroimaging Initiative: A scalable and rigorous approach to multisite amyloid and tau quantification. Alzheimer’s & Dementia 21(1), 14378 (2025) 10.1002/alz.14378

[9] Hao, X., Li, X., Zhang, G.-Q., Tao, C., Schulz, P.E., Cui, L.: An ontology-based approach for harmonization and cross-cohort query of Alzheimer’s disease data resources. BMC Medical Informatics and Decision Making 23(Suppl 1), 151 (2023) 10.1186/s12911-023-02250-z

[10] Birkenbihl, C., Emon, M.A., Vrooman, H., Westwood, S., Lovestone, S., Hofmann-Apitius, M., Fröhlich, H.: Differences in cohort study data affect external validation of artificial intelligence models for predictive diagnostics of dementia - lessons for translation into clinical practice. EPMA Journal 11(3), 367–376 (2020) 10.1007/s13167-020-00216-z

[11] Lin, Y.-C., Chiu, S.-C., Massey, R., Fang, Y.-H.D.: Classification of tau status with machine learning models in amyloid-positive cohorts. Alzheimer’s & Dementia 22(8), 71683 (2026) 10.1002/alz.71683

[12] Arnold, D., Machado, L.S., Rahmouni, N., et al.: Machine learning approach for predicting amyloid and tau positivity in Alzheimer’s disease using clinically accessible features. Alzheimer’s & Dementia 21, 105937 (2025) 10.1002/alz70856_105937

[13] Brown, C.A., Mundada, N.S., Cousins, K.A.Q., et al.: Evaluation of copathology and clinical trajectories in individuals with tau-clinical mismatch. JAMA Neurol. 83(2), 126–136 (2026) 10.1001/jamaneurol.2025.4974

[14] LaMontagne, P.J., Benzinger, T.L.S., Morris, J.C., Keefe, S., Hornbeck, R., Xiong, C., Grant, E., Hassenstab, J., Moulder, K., Vlassenko, A., Raichle, M.E., Cruchaga, C., Marcus, D., et al.: OASIS-3: Longitudinal neuroimaging, clinical, and cognitive dataset for normal aging and Alzheimer disease. medRxiv (2019) 10.1101/2019.12.13.19014902

[15] Mishra, S., Gordon, B.A., Su, Y., Christensen, J., Friedrichsen, K., Jackson, K., Hornbeck, R., Balota, D.A., Cairns, N.J., Morris, J.C., Ances, B.M., Benzinger, T.L.S.: AV-1451 PET imaging of tau pathology in preclinical Alzheimer disease: Defining a summary measure. NeuroImage 161, 171–178 (2017) 10.1016/j.neuroimage.2017.07.050

[16] Open Access Series of Imaging Studies (OASIS): OASIS-3 Imaging Methods and Data Dictionary, Version 2.3 (Data Release 2.0). https://sites.wustl.edu/oasisbrains/home/oasis-3/. Accessed 16 October 2026 (2022)

[17] Gebre, R.K., Moscoso Rial, A., Raghavan, S., Wiste, H.J., et al.: Advancing tau PET quantification in Alzheimer disease with machine learning: Introducing THETA, a novel tau summary measure. Journal of Nuclear Medicine 65(9), 1473 (2024) 10.2967/jnumed.123.267273

